# Lack of coordination and medical disinformation in Canadian self-assessment tools for COVID-19

**DOI:** 10.1101/2020.04.14.20065631

**Authors:** Brieanne Olibris, Amir Attaran

## Abstract

As SARS-CoV-2 threatens to overwhelm health systems in Canada, it is imperative that provinces are able to plan and manage an effective and reduced risk response. For this response to be most effective, it must reflect an evidence-based, pan-Canadian response. We designed four different prototypical patients with a combination of common COVID-19 symptoms and opportunities for exposure who were made to self-assess using the 10 provincial COVID-19 self-assessment tools on 1 April. These tools were developed to allow individuals to self-triage, allowing health systems direct capacity to testing and care. We assessed the consistency of the self-assessment tools and of the guidance provided to the patients. While the tools generally screen in three areas, the scope of included COVID-19 associated symptoms as well as the opportunities for exposure, and therefore transmission, vary between provinces such that no two provinces screened in the same way. This was, in turn, reflected in the inconsistency in guidance found. A patient with cough who had travelled abroad or had close contact with a confirmed case within 14 days received the most consistent guidance, with remaining patients receiving guidance ranging from mandatory quarantine or self-isolation to being told they did not have COVID-19 symptoms, guidance at odds with medical evidence. Thus, there is not a single, evidence-based Canadian standard of care simply for self-assessment. Without consistency in public health guidance, Canadians cannot appropriately self-isolate to mitigate community transmission, nor can the necessary valid and reliable data be collected to inform critical epidemiological models that help guide pandemic response. If federal and provincial governments are unable to coordinate a response, Parliament must use its available jurisdiction to legislate a duty on both to follow national standards, so as to improve coordination on COVID-19 in coming months.

## Introduction

The rapid emergence of SARS-CoV-2 as a pandemic pathogen threatens to overwhelm health systems in Canada, from primary to tertiary care levels. Canadian provinces have developed web- and mobile app-based self-assessment tools to help individuals assess whether they need testing for the virus and/or medical care. The intent is that if individuals, guided by easy to use tools, are themselves able to triage based on exposure history, severity or kind of symptoms, and relevant comorbidities, the capacity of the health system for both testing and treatment is more efficiently allocated to the most urgent cases, and the risk of nosocomial transmission to uninfected persons in health care facilities is reduced.

But for this strategy to work, there are certain preconditions. Chief among these is that the self-assessment tools offered by the provinces be evidence-based and therefore result in clinically appropriate triage. Connected to this is the expectation that at any given moment in time the decision algorithm that lies behind the self-assessment tools (usually a questionnaire) is consistent across all provinces, reflecting a single, pan-Canadian standard of care, save for rare instances where necessity might force a region to adopt a lower standard of care. Consistency is also necessary to generate meaningful, “apples-to-apples” epidemiological comparisons, because if different provinces’ tools employ different decision algorithms for who should undergo SARS-CoV-2 testing, then the resulting data on COVID-19 confirmed case incidence and pattern of viral transmission will contain systematic errors between provinces, even assuming that the testing technology used is identical.

In this study, we examine the consistency of the provinces’ self-assessment tools and algorithms. We do so by experimentally testing each tool against four prototypical but hypothetical “patients” (no actual patient data was used). The output of this experiment demonstrates where the tools and algorithms possess consistency or inconsistency, both in their inputs that are visible to the user, and in the outputs that result and which are intended to guide patient behavior and in turn clinical care.

## Methods

We designed four different prototypical patients, entering self-assessment with different symptoms that may be prognostic of COVID-19. Each patient was designed based on information from the Public Health Agency of Canada (PHAC) in the week prior to 1 April, which identified cough, chills and fever as the three most frequently reported symptoms as of 26 March (1), cough, headache and chills as the three most frequently reported symptoms from 27-30 March (2-5), and cough, headache and general weakness thereafter. (6) Since it is unlikely and perhaps unreasonable to expect provinces to update screening tools daily, we used cough, chills and headache in the construction of our prototypical patients. The patients are as follows:

- Patient A: has a cough and *<u>has either</u>* travelled outside Canada *<u>or</u>* had contact with a confirmed case in the 14 days prior to symptom onset.
- Patient B: has a cough and has *<u>no</u>* history of travel outside Canada nor any contact with a confirmed case in the 14 days prior to symptom onset.
- Patient C: has a headache and chills and *<u>has either</u>* travelled outside Canada *<u>or</u>* had contact with a confirmed case in the 14 days prior to symptom onset.
- Patient D: has a headache and chills and has *<u>no</u>* history of travel outside Canada nor any contact with a confirmed case in the 14 days prior to symptom onset.

For Patients A and C, we merged contact with a confirmed case and travel outside Canada, not because they are causally similar or *a priori* give rise to comparable risks, but because all provinces assess both as risk factors for exposure and infection, making separation unnecessary. Each of these patients was made to self-assess on 1 April 2020, using the tools provided by each province on that date. (7-16) In each iteration, we noted which questions were presented to the patient by the tools and recorded the medical guidance that the algorithm provided the patient in the final result. This method results in two categories of reportable findings: (1) whether the tool itself posed questions that are consistent with other provinces, and; (2) whether patients received consistent medical guidance across the provinces.

## Results

### Consistency of self-assessment tools

Generally the self-assessment tools can be described as having three parts: (i) screening for health events that require immediate or urgent medical attention, (ii) screening for COVID-19 associated symptoms, and (iii) screening for possible exposure, either through travel outside Canada or contact with a confirmed case. For clarity of presentation we present results for each of these parts in turn.

With respect to Part One: Seven out of ten provinces (British Columbia, Alberta, Saskatchewan, Manitoba, Ontario, Newfoundland, Prince Edward Island) begin with a variation on a question such as this one, which is quoted from the British Columbia tool (7):

> *Are you experiencing any of the following:*
>
> *Severe difficulty breathing (e*.*g. struggling to breathe or speaking in single words)*
>
> *Severe chest pain*
>
> *Having a very hard time waking up Feeling confused*
>
> *Losing consciousness*

Minor variations of this question exist, for example Manitoba’s focus on ‘extreme drowsiness’ rather than difficulty awakening, and Ontario’s omission of either. Prince Edward Island presents a more extensive list, including headache and child-specific symptoms, though not every question is related to COVID-19 (e.g. injury requiring sutures or indicative of a broken bone).

Three provinces (British Columbia, Alberta and Saskatchewan) take this line of questioning further, following-up with a variation of the question below:

> *Are you experiencing any of the following:*
>
> *Mild to moderate shortness of breath*
>
> *Inability to lie down because of difficulty breathing*
>
> *Chronic health conditions that you are having difficulty managing because of difficulty breathing* (7)

Manitoba asks a similar question but omits inquiries about chronic health conditions until later in its tool, while Ontario seeks information about chronic conditions and immune health. For all seven provinces, there is a clear priority to identify any symptoms requiring immediate or urgent medical attention first, in order to advise respondents to seek appropriate medical attention without having them proceed through any other screening questions.

With respect to Part Two: All provinces inquire into COVID-19 related symptoms, but the scope of questions posed in their tools varies greatly from narrow to broad. Saskatchewan and Newfoundland & Labrador only screen for fever, cough and shortness of breath (or difficulty breathing for New Brunswick). British Columbia, Alberta, Manitoba, Ontario, Quebec and Prince Edward Island start with this narrow base, but more broadly incorporate additional symptoms: in no particular order, sore throat, fatigue, headache, muscle aches, sneezing, runny nose, and lethargy or poor feeding in infants and young children. Notably, Quebec is the only province to integrate screening for anosmia into their tool. A full comparison of screened symptoms by province is available in Table 1.

**Table 1.** Symptoms included in self-assessment screening by province.

| Symptom | Province |  |  |  |  |  |  |  |  |  |
| --- | --- | --- | --- | --- | --- | --- | --- | --- | --- | --- |
|  | BC | AB | SK | MB | ON | QB | NB | PE | NS | NL |
| Cough | ✓ | ✓ | ✓ | ✓ | ✓ | ✓ | ✓ | ✓ | ✓* | ✓ |
| Fever | ✓ | ✓ | ✓ | ✓ | ✓ | ✓ | ✓ | ✓ | ✓* | ✓ |
| Shortness of breath |  | ✓ | ✓ | ✓ | ✓ | ✓ |  |  |  | ✓ |
| Sore throat | ✓ | ✓ |  |  | ✓ |  |  |  |  |  |
| Runny nose |  | ✓ |  | ✓ | ✓ |  |  | ✓ |  |  |
| Sneezing | ✓ |  |  |  |  |  |  | ✓ |  |  |
| Muscle aches/myalgia |  |  |  |  |  |  |  |  |  | ✓ |
| Fatigue |  |  |  |  | ✓ | ✓ |  | ✓ |  |  |
| Headache |  |  |  |  | ✓ |  |  |  |  |  |
| Difficulty breathing |  | ✓ |  |  |  |  | ✓ |  |  |  |
| Sudden loss of smell |  |  |  |  |  | ✓ |  |  |  |  |
| Poor feeding and lethargy in infant |  |  |  | ✓ | ✓ |  |  |  |  |  |

\*-In Nova Scotia, self-assessment tool respondents are only asked about symptoms if they answer 'Yes' to initial screening questions about recent travel history or close contact with a confirmed case or a returned traveler showing respiratory symptoms.

Two provinces are exceptions to this general format of symptom inquiry. New Brunswick’s tool is unique because it does not contain a decision algorithm, but rather a list of bespoke situations, each a different permutation of symptoms, travel history, contacts, comorbidities, employment, and/or residence. When a patient fits one of these situations, there is advice to seek medical care (immediately, or if less serious, soon) or to stay at home, but a patient outside those situations is given no advice, which is true of Patient B (if under 60 years of age and not a health care worker). Nova Scotia’s tool does not ask respondents for information about their symptoms unless they have returned to the province within the 14 days prior to using the tool or unless they indicate that they have had contact with a confirmed case or someone who is a possible case, defined in Nova Scotia as someone who has returned from out of province or country with a fever or cough. A critical deficiency of Nova Scotia’s tool is that it can prematurely draw conclusions without input as to the patient’s symptoms, but rather just the travel history of the patient or the travel history and symptoms of others.

With respect to Part Three: All ten provinces screen respondents for potential exposure to the virus, but the opportunities for exposure, and therefore transmission, are varied. With the exception of Quebec, all provinces screen for travel outside of Canada and/or outside of the province within 14 days of using the tool or of symptom onset, which is consistent with the early (and now obsolete) perspective that COVID-19 is a foreign threat.

All provinces except Alberta screen for exposure to a confirmed case. The exposure in this situation is explicitly defined as or implied to be close contact, though Manitoba’s tool screens for multiple levels of proximity to a confirmed case. Saskatchewan and Manitoba also screen for exposure via contact with a case under investigation (awaiting test results). British Columbia, Manitoba, Ontario, Newfoundland & Labrador, New Brunswick, Nova Scotia and Prince Edward Island also screen for close contact with a possible case, but there is no agreement between the provinces on the definition of possibility. For example, British Columbia provides no definition, while in Ontario, a possible case is any individual who has new respiratory symptoms or anyone who has “recently” travelled outside of Canada, and five provinces (Manitoba, Newfoundland & Labrador, New Brunswick, Nova Scotia, and Prince Edward Island) define a possible case as an individual who travelled outside of the province and either has “respiratory symptoms”, “respiratory illness” or cough and/or fever. Further, how the latter five provinces measure time differs: New Brunswick inquires whether the user has travelled and returned within 14 days of symptom onset, while the remaining provinces count 14 days from the possible case’s symptom onset which in effect neglects the possibility of community transmission.

Tools also vary in terms of whether the emphasis is placed on exposure (and thus these are the first questions in the decision tree) or symptoms (likewise placed first in the decision tree), and the relative importance given to the two in the tool. We identified no obvious superiority in either approach.

In addition to the self-assessment tool for the general public, British Columbia (17) and Alberta (18) both have specific tools or COVID-19 testing criteria for health care workers. In British Columbia, for health care workers the symptoms which trigger COVID-19 testing are broader than the public (including diarrhea, fatigue and rhinorrhea). In Alberta, the symptoms are the same, but health care workers are not asked about their travel history. The reasons for these discrepancies are unknown.

### Consistency of guidance

The greatest consistency in guidance is provided for our Patient A, who has a cough and has either travelled outside of Canada in the 14 days prior to symptom onset or has had contact with a confirmed case. Every province except Quebec explicitly instructs the respondent to self-isolate for 10-14 days (the duration varies by province) from symptom onset in either scenario, a recommendation which is layered with the legal requirement for 14 days self-quarantine by travelers returning to Canada. (19) If Patient A lives in the provinces screening for contact with possible cases (discussed in the previous section) they will be provided with the same guidance.

All provinces advise Patient A to contact a health information line or a primary care provider based on identifying with a cough, except British Columbia which advises Patient A to make contact only if symptoms worsen or Patient A is concerned. In Quebec, an individual can identify if there is a confirmed case in their immediate circle but not whether they have travelled, such that if Patient A travelled they are advised to follow prevention measures for the general public (e.g. physical distancing), but if Patient A has had contact with a “confirmed case of COVID-19 with a laboratory test” they are advised they may need screening and provided with information to book an appointment for COVID-19 testing. (12) By inquiring only about confirmed, and not possible, cases, Quebec’s tool makes the accuracy of a Patient A’s triage dependent on the speed of testing his or her contacts.

Patient B, who also has a cough but has not travelled outside of Canada or had contact with a confirmed case is sometimes asked to self-isolate for 10-14 days (the duration varies by province) after symptom onset, similar to Patient A. For example, Manitoba and Nova Scotia advise anyone with symptoms to self-isolate until symptoms have resolved. In Quebec, Patient B receives the same advice as Patient A who has travelled. In New Brunswick, the tool does not offer the user options corresponding to Patient B, who consequently does not receive guidance. In Saskatchewan, Patient B is simply told to self-monitor. Finally, in Newfoundland & Labrador, Patient B provided with the following guidance:

> *There are many common viruses other than COVID-19 that can cause your symptoms. Based on your responses you do not need to be tested for COVID-19 at this time*.
>
> *In general, if you are experiencing cold or flu-like symptoms, you should practice good cough and sneeze etiquette and stay at home until your symptoms resolve*. (16)

In Alberta, if Patients A or B are health care workers, they would be told that testing may be necessary, though Patient B in the general public is informed COVID-19 testing is not needed.

Notably, while Patients A and B present with cough, the above results would be identical in all provinces if presenting with fever instead.

Based on symptoms alone, Patients C and D are the most likely to be missed in screening, despite headache and chills being common symptoms, more so than other symptoms included by some provinces. No province’s tool screens for chills. Only Ontario and PEI’s tools screen for headache. In PEI, Patients C and D will be directed to call 911 or attend an emergency department for persistent headache.

Contrariwise in British Columbia, Saskatchewan and Newfoundland & Labrador patients C and D are given dangerous disinformation that they “don’t have any COVID-19 symptoms” and do not need to be tested. (7,9,16) Otherwise, Patient C and patient A are given similar advice, due to travel.

## Discussion

All Canadian provinces have developed COVID-19 self-assessment tools that allow the general public to assess what action they should take based on symptoms, exposure to others, and travel. However, the similarities end there as no two provinces engage in the same screening. Even in a cross-Canada pandemic as devastating as this, there is not a single, evidence-based Canadian standard of care simply for self-assessment. This is despite the Public Health Agency of Canada publishing a national COVID-19 case definition on February 6, 2020 (20), since updated from time to time to be consistent with WHO definitions. (21)

Instead each province has devised a tool, based its own case definition or perception of the scientific evidence, inevitably resulting in chaos and errors. To use a metaphor, not only have provinces wasted resources and time “reinventing the wheel”, but some of their wheels are more oval than round.

Most concerning is the fact that some provinces have developed their tools at variance with medical evidence, and these tools negligently furnish disinformation on a potentially deadly illness.

It is likely that provinces used sources of information from outbreaks in other countries to formulate their decision algorithms. The Report of the WHO-China Joint Mission on Coronavirus Disease 2019 (COVID-19), released in February 2020 lists these diagnostic symptoms in descending order: fever, dry cough, fatigue, sputum production, shortness of breath, sore throat, headache, myalgia or arthralgia, chills, nausea or vomiting, nasal congestion, diarrhea, hemoptysis and conjunctival congestion. (22) Similar advice is given by the Public Health Agency of Canada in its daily epidemiology updates, and at the end of March 2020, cough, headache and chills were the three most frequently reported symptoms. (6)

Yet many provinces have created self-assessment tools that ignore this medical evidence. Most alarming is British Columbia, Saskatchewan and Newfoundland & Labrador informing users with headache and chills (Patients C and D) that they do not have COVID-19 symptoms—negligent advice that were it given by a clinician rather than a computer would be medical malpractice.

Similarly concerning is Nova Scotia’s tool, which considers only travel-associated risks and entirely overlooks that COVID-19 can be transmitted in the province without travel. Perhaps aware of this failure Nova Scotia withdrew its self-assessment tool after the data for this study was collected, and as of this writing (8 April 2020), it has been replaced with a single screen advising individuals with certain symptoms to call a helpline for further assessment.

There are two extremely serious epidemiological consequences to the failure of the provinces to align on a single standard of care in self-assessment. First, without consistent public health advice as to who has COVID-19 symptoms, or agreement on what constitutes a possible COVID-19 case, there is a fundamental failure to ensure that Canadians appropriately self-isolate to mitigate community transmission, which risks causing infection and illness to others and worsening the pandemic. Second, there is a fundamental failure to give appropriate advice to users about when to present for COVID-19 testing, so as to become tallied as a confirmed case if positive. This failure in turn introduces systematic errors into provincial counts of confirmed cases, making epidemiological comparisons between provinces inaccurate and misleading, and degrading the quality of critical epidemiological models—which Canada requires to guide safe de-escalation of social distancing, timely reinstatement of civic institutions and the economic activity, and overall a return to normalcy.

These failures could be avoided. Following the 2003 SARS outbreak, the federal government and provinces collaborated in assessing “lessons learned” and agreed to coordinate outbreak response. (23) The failure to have a single, federal self-assessment tool—or multiple provincial tools all conforming to a single standard of care—is proof that in 17 years, coordination has failed.

In our opinion, this failure mandates two federal responses.

In the short term (under a week), the federal government should develop a single self-assessment tool for COVID-19, in both official languages, and perhaps also languages of immigrants, which provinces should assist to develop and adopt. This would solve the immediate problem of lacking a single, evidence-based standard of care for self-assessment.

In the intermediate term (a month or two), the federal government must introduce legislation to ensure that other dimensions of the COVID-19 response are not undermined by poor coordination. As the federal government concluded in its “lessons learned” report from SARS 2003:

> *In the event that a coordinated system of rules for infectious disease surveillance and outbreak management cannot be established by the combined effects of the [federal, provincial and territorial governments] and the above-referenced intergovernmental legislative review, the*
>
> *Government of Canada should initiate the drafting of default legislation to set up such a system of rules, clarifying F/P/T interactions as regards public health matters with specific reference to infectious diseases*.*(21, p*.*216)*

The absence of a single, evidence-based standard of care for self-assessment furnishes clear evidence that, in the 17 years since SARS, provincial and federal governments cannot coordinate in the timely manner demanded by epidemics or pandemics. Parliament therefore must use its available jurisdiction to legislate a duty on both to follow national standards, so as to improve coordination on COVID-19 in coming months. (24)

## Data Availability

All tools referred to in the manuscript are publicly available. As they are subject to change, they have been archived on Wayback Machine.

